# Gait Adaptations to Walking Speeds in Individuals with Myotonic Dystrophy Type 1

**DOI:** 10.1101/2024.08.28.24312607

**Authors:** Barthélémy Hoerter, Laurent Ballaz, Yosra Cherni

## Abstract

**Background:** Myotonic dystrophy type 1 (DM1) is a prevalent inherited muscular dystrophy in adults, affecting distal muscles such as the gastrocnemius, soleus, and tibialis anterior. This leads to significant gait deviations and reduced walking speed, impacting overall well-being and increasing fall risk.

**Objective:** This study aimed to assess how walking speed affects gait kinematics in individuals with DM1.

**Method:** Eighteen individuals with genetically confirmed DM1 (4 women, age: 41.0 [35.5; 47.8] years, mass: 76.8 [67.1; 94.6] kg, height: 166.0 [156.7; 173.3] cm) participated in this study. Each participant walked barefoot along a 13-meter walkway at comfortable and fast speeds. Subsequently, spatiotemporal parameters and joint kinematics were assessed.

**Results:** The step length (p < 0.001), cycle speed (p < 0.001), and cadence (p < 0.001) increased significantly, leading to a higher walking speed. Moreover, the vertical amplitude of the center of mass (CoM) increased significantly (p = 0.015), while the mediolateral amplitude decreased (p = 0.001) at fast walking condition. In addition, significant kinematic changes included increased trunk tilt (p < 0.001), greater anterior pelvic tilt (p < 0.001), increased hip flexion at initial contact, and enhanced knee flexion during both stance and swing phases. Ankle dorsiflexion showed a trend towards increase during stance phase (p = 0.055) at fast walking condition.

**Conclusions:** Fast walking speed in individuals with DM1 lead to significant gait adaptations. These adaptations reflect compensatory mechanisms to manage muscle weakness. The present study revealed significant changes in spatiotemporal parameters related to walking speed. Fast walking also highlighted kinematic adaptations in trunk, pelvis and lower limb joints. These findings enhance our understanding of gait deviation in individuals with DM1 and suggest the potential benefits of targeted fast walking training in this population.

## 1. INTRODUCTION

Myotonic dystrophy type 1 (DM1) is the most prevalent inherited muscular dystrophy in adults [1], affecting 3-15 per 100.000 people over Europe. However, its prevalence is notably higher in specific areas such as the province of Quebec (Canada), affecting around 1 individual per 500 [2]. Clinically, DM1 is characterized by progressive weakness and myotonia of distal muscles, as well as cardiomyopathies, central nervous system dysfunctions, and decreased visual acuity [3–5]. Muscle contraction and relaxation delays are significantly prolonged [4,6], and some muscles exhibit abnormal activation [7], leading to excessive and early fatigue during daily activities, such as walking [8,9].

These muscle disorders in individuals with DM1 result in notable gait deviations, altered coordination, and a reduced motor repertoire [10,11]. Gait deviations in individuals with DM1 are mainly manifested by a reduced walking speed, cadence, stride length, and joint range of motion (RoM), along with increased stance phase duration in comparison to typically developing individuals [12]. Consequently, gait and balance disorders are prominent in individuals with DM1, leading to an increased risk of falls [13]. Given the essential role of walking in maintaining autonomy and independence, the ability to perform daily tasks is reduced in individuals with DM1 [14]. Walking speed is directly linked to a person’s available energy, where slower gait indicates lower energy and vice versa. Indeed, regression analysis has demonstrated a negative correlation between available energy and age, implying that walking speed is an indicator of frailty and disability [15]. Furthermore, walking speed is recognized as an indicator of autonomy, with speeds below 0.8 m/s indicating disability [16,17]. The impact of reduced walking speed in individuals with DM1 is further highlighted by Hammarén et al. (2015), which observed a significant decrease in walking speed over five years in individuals with DM1, indicating a rapid decline in walking capacity. The loss of this walking capacity is primarily due to a rapid decrease in the maximal isometric strength of distal muscles, particularly the ankle dorsiflexors, which showed a 12% reduction in strength over five years [18]. Again, Filli et al. (2020) and Tiffreau et al. (2018) each examined the gait kinematics of individuals with DM1 by having participants walk on a treadmill and comparing them to typically developing individuals [6,19]. In Filli et al. (2020)’study, the selected walking speeds were 1 km/h and 2.5 km/h, both within a comfortable range. They observed a significant decrease in ankle dorsiflexion during the swing phase and a trend towards decreased step length with increasing speed in individuals with DM1 compared to typically developing individuals [19]. Tiffreau et al. (2018) observed shorter step length in individuals with DM1 compared to typically developing individuals [6]. This was compensated by an increase in cadence to adapt to the treadmill speed. It has been suggested that ankle plantar flexor weakness and abnormal muscle co-activations, due to the prolonged gastrocnemius contraction, limit the ability to lengthen the step [6]. However, these studies have mainly focused on walking at comfortable or treadmill-controlled speeds, which do not accurately reflect the short periods of rapid walking necessary in daily life, such as when crossing a street.

The aim of this study was to evaluate the effect of different walking speeds on spatiotemporal and kinematic parameters in individuals with DM1. At fast walking speed, individuals with DM1 are expected to show impaired movement more pronounced at the ankle due to the weakness of distal muscles such as the gastrocnemius and the tibialis anterior, the latter of which can cause foot drop. Furthermore, we anticipate that the increase in walking speed will result in compensatory mechanisms such as increased step cadence and shorter step length.

## 2. MATERIAL AND METHODS

### 2.1 Participants

The inclusion criteria, which were assessed by a clinician, were: (1) a genetically confirmed diagnosis of DM1, (2) being at least 20 years old, (3) being able to walk with or without assistive device, and (4) to demonstrate comprehension and communication abilities. The exclusion criteria were: (1) having undergone physical training during the last 12 months, (2) having had surgery in the 12 months prior to the study, and (3) having a contraindication to physical activity. This project received approval from the ethics committee of the Centre de Recherche Interdisciplinaire en Adaptation Réadaptation (Montreal, Canada). All participants provided their informed consent before participation.

### 2.2. Experimental protocol

#### a. Kinematics

An optoelectronic system (Vicon T40Sx, Oxford, UK) with a 12-camera, operating at 100 Hz, was used to capture marker trajectories during trials. Thirty-seven reflective markers were used according to the Plug-in Gait kinematic model [20]. A 13-meter walkway was prepared, where participants walked barefoot along the walkway at a (1) self-selected comfortable speed and then (2) at their maximum achievable walking speed without turning into running. For each walking condition, a minimum of three trials were recorded with 30-second rest intervals in between.

#### b. Muscle Strength

Maximal isometric strength of the knee and hip flexors and extensors, as well as hip abductors were measured using a handheld dynamometer (Lafayette manual muscle test system, Lafayette, USA). According to the protocol outlined by Eek et al. (2008), the knee flexor and extensor strength was measured in a sitting position, hip flexor and abductor strength in a supine position, and hip extensor strength in a prone position [21]. During the measurements, the assessor positioned the device on the distal part of the thigh or shank, while an assistant stabilized the proximal segment. Participants were instructed to exert maximum effort for 6 seconds. Three trials were conducted for each muscle group, with a 30-second rest period between each trial. Finally, the maximal isometric strength was calculated by multiplying the measured force in dynamometry by the lever arm (Nm). Maximal isometric strengths are listed in Table 1.

**Table 1:** Characteristics of participants and maximal isometric strength of lower limb muscle groups (M: Men; F: Women; DM1: myotonic dystrophy type 1).

| Participants characteristics |  |
| --- | --- |
| Sex | 4F; 14 M |
| DM1 type | 9 Adult-onset; 9 Congenital |
| age (years) | 41 [36; 48] |
| Mass (kg) | 76.8 [67.1; 94.6] |
| Height (cm) | 166.0 [156.7; 173.3] |
| Maximal isometric strength level (Nm) |  |
| Hip flexors | 75.6 [62.1; 94.6] |
| Hip extensors | 51.2 [38.0; 76.5] |
| Hip abductors | 66.0 [53.1; 75.5] |
| Knee flexors | 45.5 [34.8; 58.0] |
| Knee extensors | 78.6 [52.2; 95.7] |

### 2.3. Data processing

Marker trajectories were processed using Nexus 2.14.0 (Vicon Motion Systems, Oxford, UK). The participants anthropometric dimensions were measured and used to calculate joint positions [22]. A Custom-made MATLAB library (vR2022a, Mathworks, Natick, USA) was used to calculate the spatiotemporal and kinematic parameters. The joint angles were calculated in all anatomical planes for the trunk, pelvis, hip, knee and ankle. Given the side-to-side symmetry in gait patterns of individuals with DM1 [7,19], right and left gait cycles were averaged for analysis. An average of 11 ± 4 cycles per participants were analyzed for self-selected comfortable walking speed and for fast walking.

### 2.4. Statistical analysis

The normality of the spatiotemporal parameters was assessed using the Kolmogorov-Smirnov test [23]. Based on the normality results, the Wilcoxon test was used to assess the effect of walking speed on spatiotemporal parameters. The results were reported as median [Q1; Q3]. Consequently, the effect size was calculated using the correlation coefficient (r) for non-parametrical tests based on the Wilcoxon Z score [24]. The normality of the time series kinematic data was tested using the D’Agostino-Pearson K2 test. Subsequently, the different kinematic curves of comfortable and fast walking were compared using Statistical Parametric Mapping (SPM) with the non-parametric SPM1D package [25]. Only the areas of significance covering at least 5% during the gait cycle were retained for our analysis [26]. For the areas of significance, the effect size was calculated using the Cohen’s d formula [24]. A statistically significant difference was accepted as p < 0.05, and the effect size was interpreted based on Cohen’s d guidelines, with 0.20 considered small, 0.50 as moderate, and 0.80 large. All the statistical analysis were performed using Matlab software (vR2022a, Mathworks, Natick, USA).

## 3. RESULTS

### 3.1. Participants

Eighteen participants with DM1 were included in this study. Participants’ characteristics are displayed in Table 1. There was an equal distribution of participants with congenital and adult-onset DM1. The participant group consisted of 4 women and 14 men, with an age ranged from 20 to 56 years. Isometric muscle strength levels were reported in Table 1.

#### a. Spatiotemporal parameters and center of mass displacement

Spatiotemporal and CoM parameters are reported in Table 2. When asked to walk faster, the participants increased significantly their walking speed by 42% (p < 0.001). The increase in walking speed was due to an increase in both step length (p < 0.001, r = 0.878) and cadence (p < 0.001, r = 0.878). The double support time decreased significantly at fast walking condition (p < 0.001, r = 0.878). However, the maximum support base width did not show any significant change related to walking speed (p = 0.983). The maximum height of the CoM did not change significantly between the two walking speeds (p = 0.238). The vertical amplitude of the CoM, however, increased significantly during fast walking (p = 0.015, r = 0.621), and the mediolateral amplitude of the CoM decreased significantly during fast walking (p = 0.001, r = 0.816).

**Table 2:** Spatiotemporal parameters in participants with DM1 during comfortable walking and fast walking.

|  | Comfortable speed | Fast speed | p-value | Effect size |
| --- | --- | --- | --- | --- |
| Walking speed (m/s) | 0.95 [0.81; 1.03] | 1.35 [1.19; 1.49] | < 0.001 | 0.878 |
| Step length (m) | 0.51 [0.47; 0.55] | 0.60 [0.53; 0.69] | < 0.001 | 0.878 |
| Cadence (number of steps/min) | 1.78 [1.63; 1.87] | 2.13 [1.97; 2.22] | < 0.001 | 0.878 |
| Double support time (% of cycle) | 35.80 [31.90; 39.80] | 27.60 [22.90; 30.35] | < 0.001 | -0.878 |
| Mean CoM height (% of height) | 55.35 [54.40; 56.13] | 54.70 [53.95; 55.55] | 0.001 | -0.785 |
| Max CoM height (% of height) | 56.35 [55.53; 57.075] | 56.10 [55.30; 57.23] | 0.238 | 0.231 |
| Vertical CoM amplitude (% of height) | 2.05 [1.58; 2.50] | 2.35 [1.85; 3.35] | 0.015 | 0.621 |
| Mediolateral CoM amplitude (mm) | 56.67 [44.81; 67.25] | 38.35 [33.6325; 54.3075] | 0.001 | -0.816 |
| Max support base width (mm) | 202.56 [169.18; 264.98] | 219.53 [163.17; 264.95] | 0.983 | 0.010 |

#### b. Kinematic parameters

The joint angles in sagittal plane were calculated at specific moments and are displayed in Table 3. The maximum anterior trunk tilt increased significantly during the stance phase while walking fast (p = 0.005, r = 0.667). The average anterior pelvic tilt during the stance phase increased (p = 0.001, r = 0.755). The hip flexion angle at initial contact increased significantly (p = 0.001, r = 0.796), and the hip flexion angle at toe-off decreased (p < 0.001, r = 0.867). Maximal knee flexion significantly increased during single limb stance at fast walking (p < 0.001, r = 0.847), and the mean CoM height decreased (p = 0.001, r = -0.785). The ankle angle showed a trend towards increased dorsiflexion (p = 0.055, r = 0.452).

**Table 3:** Pelvic, hip, knee, and ankle angles in participants with DM1 during comfortable walking and fast walking.

| Angle in the sagittal plane (deg) | Comfortable speed | Fast speed | p-value | Effect size |
| --- | --- | --- | --- | --- |
| Max. trunk tilt during stance phase | 7.04 [3.56; 14.73] | 8.78 [6.02; 20.20] | 0.005 | 0.667 |
| Mean pelvic tilt during stance phase | 16.27 [9.89; 18.36] | 16.74 [13.41; 19.04] | 0.001 | 0.755 |
| Mean hip flexion at initial contact | 37.10 [29.86; 41.00] | 42.65 [36.55; 46.54] | 0.001 | 0.796 |
| Mean hip angle at push-off | 6.90 [1.21; 13.80] | 0.84 [-4.17; 8.89] | < 0.001 | -0.867 |
| Max. knee flexion during single limb stance | 17.5 [8.89; 25.69] | 23.41 [18.87; 32.67] | < 0.001 | 0.847 |
| Max. ankle dorsiflexion during single limb stance | 15.75 [11.00; 17.88] | 16.88 [12.55; 20.32] | 0.055 | 0.452 |

The joints kinematics during gait was recorded in the three anatomical planes and are reported in Figure 1. **In the sagittal plane**, the trunk kinematics demonstrated higher tilt during fast walking throughout the entire gait cycle (p < 0.001, d = 0.251). An increase in anterior pelvic tilt was noted throughout the entire gait cycle (p < 0.001, d = 0.237). At hip joint, an increase in flexion was observed between 0% and 34% of gait cycle (p < 0.001, d = 0.307). Moreover, an increase in hip extension was observed between 45% and 58% of the gait cycle (p = 0.001, d = -0.201). Regarding the knee joint, an increase in flexion was observed at the beginning of the gait cycle, from 0% to 34% (p < 0.001, d = 0.370). Knee flexion also increased significantly during the swing phase, with a medium effect, from 54% to 85% of the gait cycle (p < 0.001, d = 0.544). Finally, a significant increase in ankle dorsiflexion was noted during single limb support, from 11% to 48% of the gait cycle (p < 0.001, d = 0.452).

**Figure 1:**
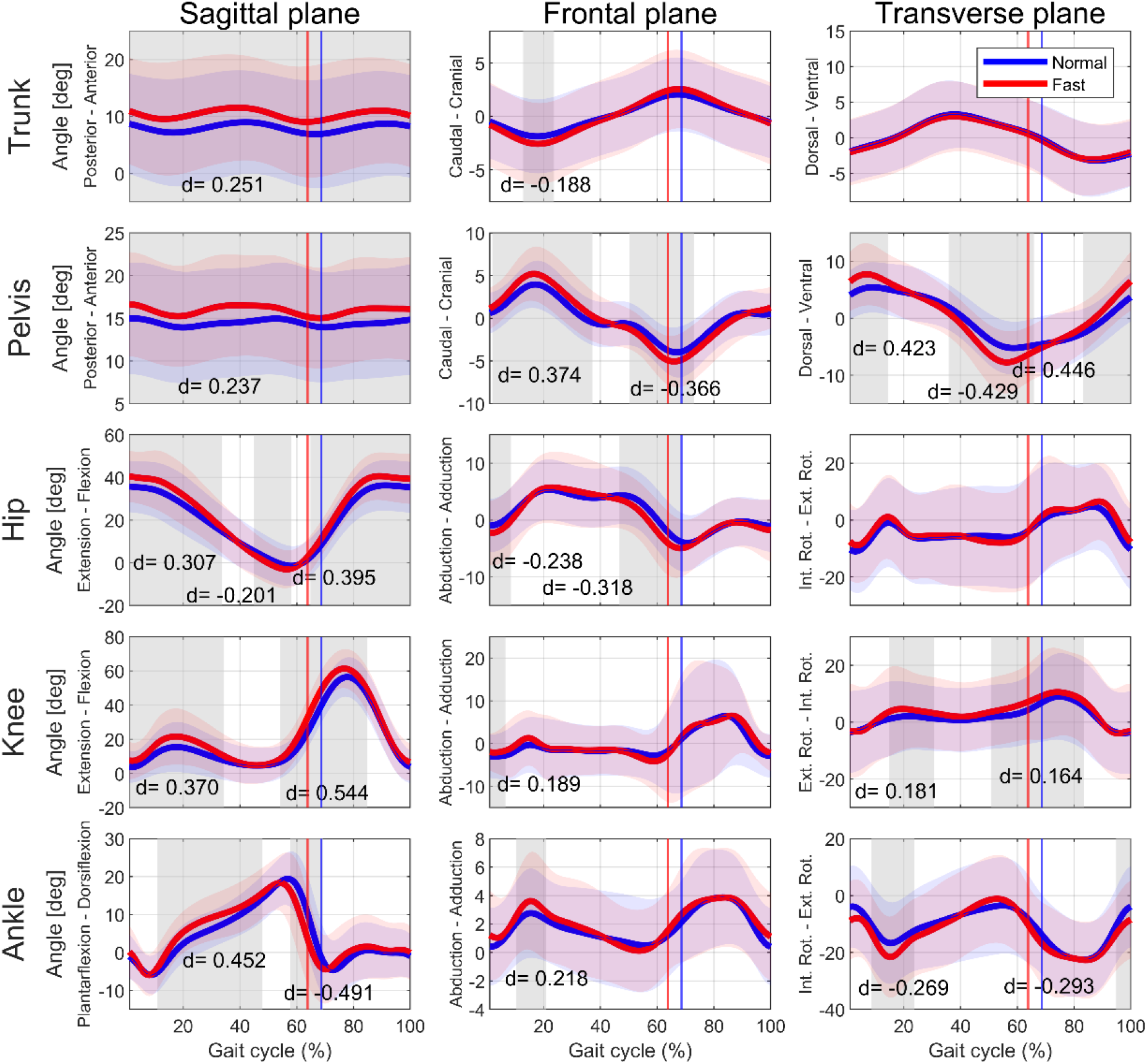
Trunk, pelvic, hip, knee and ankle angles in the sagittal, frontal and transverse planes for comfortable walking and fast walking. The shaded areas represent significant differences between the two curves according to SPM1D. Cohen’s d values were calculated for each shaded area.

Regarding **the frontal plane**, a significant lateral tilt of the trunk was observed during the stance phase, on the ipsilateral side of the supporting leg (p = 0.011, d = -0.188). A higher lateral pelvic obliquity was shown during fast walking between 2% and 37% of the gait cycle as well as between 50% and 74% of the gait cycle (p < 0.001, d = 0.374 and p < 0.001, d = -0.366, respectively). Hip abduction during initial swing was consistent with the pelvic obliquity, showing a small effect (p < 0.001, d = -0.318).

**In the transverse plane**, no difference was observed at the trunk rotation. However, a greater pelvic rotation was noticed during fast walking compared to comfortable walking (p = 0.001 and d = 0.423, p < 0.001 and d = -0.429, p < 0.001 and d = 0.446 depending on the gait cycle phase). No significant change was observed at the hip rotation.

## 4. DISCUSSION

The objective of this study was to evaluate the effect of walking speed on kinematic parameters in individuals with DM1. Our findings revealed that during fast walking, individuals with DM1 exhibited a significant increase in step length and cadence. Additionally, kinematic analysis indicated an increase in anterior pelvic tilt, hip flexion and extension, knee flexion, and ankle dorsiflexion, as well as greater pelvic obliquity and rotation.

### a. Impact of walking speed on spatiotemporal parameters

Several parameters contributed to the increased walking speed in individuals with DM1 such as an increase in cadence, step length, as well as a decrease in double support time. The increase in cadence aligns with the observations of Tiffreau et al. (2018), who noted this mechanism to adapt to walking speed on a treadmill in individuals with DM1 [6]. Similarly, Missaoui et al. (2010) reported a trend towards increased cadence without an increase in step length as a fast walking strategy in individuals with DM1 [5]. Interestingly, some studies have reported a decrease in step length accompanied by an increase in cadence to adapt to the treadmill-imposed speed in individuals with DM1 [6,19]. In the other hand, our findings indicated that both step length and cadence increased during fast walking in individuals with DM1, which align with the results of Lim et al. (2017) in typically developing individuals [27]. This discrepancy may be attributed to the fact that we did not control walking speed using a treadmill. Indeed, some studies advise caution when comparing overground walking with treadmill walking [28]. According to the study by Azizah Mbourou et al. (2003), a decrease in step length is associated with increased risk of falling in typically developing individuals [29]. The same study also found that adults who are prone to fall spend more time in double support than non-fallers [29]. In the present study, the increase in step length was accompanied by a decrease in double support time during the stance phase, which is linked to the increase in walking speed and suggests a reduced risk of falling. Additionally, this reduction in double support time is a common mechanism to facilitate faster walking [30]. Therefore, the findings of our study suggest that faster walking speed may have a beneficial effect on fall risk in individuals with DM1. Further research is needed to explore the relationship between walking speed and fall risk in individuals with DM1, but these results indicate that training fast walking could be a beneficial aspect of intervention for this population.

### b. Impact of walking speed on kinematics

The trunk tilt, which is necessary to maintain forward movement [31,32], was greater during fast walking compared to comfortable walking in the sagittal plane, enabling participants to increase their pace more efficiently. This strategy is similar to the one used by typically developing populations [33]. This mechanism allows for the forward shift of the center of pressure, taking advantage of gravity to propel the body forward, resulting in reduced muscular effort and improved walking efficiency [32]. The anterior pelvic tilt, which is related to trunk inclination in the sagittal plane, was also higher during fast walking condition compared to comfortable walking condition. Similarly, Taylor et al. (1999) observed an association between decreased walking speed and reduced anteroposterior pelvic tilt in typically developing individuals [34]. These observations suggest that increased walking speed among individuals with DM1 is facilitated by a greater anterior pelvic tilt, reflecting a compensatory mechanism observed across different populations.

Increased lateral trunk tilt during the stance phase and greater pelvic obliquity during fast walking indicate a Duchenne limp, which suggests hip abductor weakness [35]. In individuals with DM1, a significant decline in hip abductor strength over a 3-year period has been documented by Roussel et al (2021) [36]. The hip abductor weakness is consistent with the muscle strength measurements obtained in our study which was lower than typically developing individuals. Specifically, individuals with DM1 in our study had a median maximal isometric strength of 66.0 Nm, lower than the values reported by Jacobs et al. (2005) for typically developing individuals (average strength of 81.0 ± 23.7 Nm and 76.1 ± 19.9 Nm for dominant and non-dominant leg, respectively) [37]. Some studies indicated that the general maximal isometric strength is related to functional ability [38,39]. Regarding abductor strength, Lanza et al. (2021) demonstrated a relationship between abductor strength and the Four Square Step Test, a functional performance assessment for dynamic stability in different directions [40]. Therefore, the maximal isometric strength can be used as an indicator of functional capacity in individuals with DM1. Moreover, Van Der Krogt et al. (2012) reported that hip abductor weaknesses highly impact the gait [41]. These findings highlight the importance of strengthening the hip abductors in individuals with DM1.

Regarding the hip joint, individuals with DM1 exhibited increased hip flexion at initial contact and hip extension at the end of stance phase during fast walking. These results are consistent with the findings of Fukuchi et al. (2019) in typically developing individuals [42], allowing individuals to increase their step length. Favetta et al. (2022) linked increased hip and knee flexion during the stance phase with increased anterior pelvic tilt [43]. The increase in hip RoM contributes to the observed longer stride length [44]. Additionally, the knee joint exhibited greater flexion during both the swing and stance phases in the fast walking condition, similar to that seen in typically developing individuals [45]. Increased knee flexion has been described as a shock attenuation mechanism during initial contact in typically developing individuals [46]. A similar strategy was adopted by individuals with DM1 in the present study. Indeed, the increase in walking speed necessitates greater shock attenuation, leading to increased knee flexion during initial contact and the stance phase. Lastly, the present study showed an increased ankle dorsiflexion during the stance phase in fast walking. This may be directly related to the tibialis anterior muscle prolonged activation caused by DM1 [4]. This observation is also likely due to the weakness of the gastrocnemius muscle and its delayed contraction, which fails to slow down tibial advancement during the ankle rocker phase [7]. The increase in ankle dorsiflexion also contributes to the increased hip and knee flexion during the stance phase in order to maintain upright posture [47]. These findings suggest that targeted interventions should focus on training the tibialis anterior and gastrocnemius muscles to improve lower limb joint kinematics.

### c. Impact of walking speed on CoM

An increase in the vertical CoM amplitude during fast walking compared to comfortable speed walking was observed in the present study, which was already described in typically developing individuals [48,49]. This adaptation is attributed to an increase in joint amplitudes, particularly hip and knee flexion, as observed in individuals with DM1. This mechanism, described by Mann and Hagy (1980) in typically developing individuals, is intended to lower the CoM during fast walking or running [47], which facilitates greater shock attenuation [46]. Consequently, the reduction in CoM height during shock attenuation in fast walking contributes to the observed increase in the vertical amplitude of the CoM. Furthermore, the decrease in mediolateral CoM displacement and the increase in step length are mechanisms linked to increased walking speed, as previously reported in typically developing individuals [48]. Orendurff et al. (2004) also described a decrease in step width associated with increased walking speed [48]. Contrary to these observations, we did not observe a decrease in base of support width during fast walking in individuals with DM1. Maintaining a wide step width appears to be a strategy aimed at preserving lateral stability, suggesting that individuals with DM1 have lower stability compared to typically developing individuals [50]. Dean et al. (2007) suggest that maintaining a larger step width is a way to keep the center of mass more securely within the base of support, thereby increasing confidence during walking [50]. This mechanism has been described as a strategy to counteract the sensorimotor deficits of the lower limbs, especially in older adults [51].

### d. Limitations

This study has some limitations which need to be reported. First, the number of participants was relatively small. However, the prevalence of this disease makes it challenging to gather large groups of participants. Second, the participants were the ones with congenital or adult-onset forms of DM1, which may influence the results due to the disease progressions [36,52]. However, the symptoms are similar, the main difference lies in the severity of the disease [2]. Third, the application of these study results to everyday life remains hypothetical. Further research is required to assess the validity of these measures in daily activities.

In conclusion, this study provides critical insights into how walking speed affects gait kinematics in individuals with DM1. To our knowledge, this is the first study to compare overground fast walking speed with comfortable walking speed in individuals with DM1. Significant spatiotemporal and kinematic changes were observed, which may reflect adaptive mechanisms to manage distal muscle weakness. These findings highlight gait adaptations such as increased ankle dorsiflexion during stance phase and reduced double support time.

## Data Availability

none

## ACKNOWLEDGMENTS

The authors would like to thank the participants and the physical therapists from the Centre de réadaptation Lucie-Bruneau (Montréal, Canada) for their valuable assistance. Finally, the authors would also like to thank Sina Tabeiy for reading the article for English language accuracy.

## DATA AVAILABILITY

Data supporting the results reported in the article can be provided by the corresponding author.

